# Mortality and Epidemiological Characteristics of Pulmonary Embolism in Spain (2022–2024)

**DOI:** 10.64898/2026.09.03.26362157

**Authors:** Isabel Mendo-Pedrajas, Pilar Rondon-Fernandez, Carmen de-Juan-Alvarez, Sandra Coronado-Fernández, Rafael Garcia-Carretero

## Abstract

**Background:** Acute pulmonary embolism (PE) is a major cause of cardiovascular morbidity and mortality. This study aims to describe the epidemiological trends, clinical complexity, and in-hospital mortality of PE admissions in Spain from 2022 to 2024.

**Methods:** We conducted a nationwide, retrospective study using the Spanish Minimum Basic Data Set for Hospitalization (MBDS-H) for adult patients admitted with acute PE.

**Results:** We identified 125,305 hospital admissions for acute PE. The median patient age was 74 years with a slight male predominance (51–52%). A high comorbidity burden was observed: 61–62% had COPD, 35–37% hypertension, and 21–24% malignancy; ICU admissions increased from 11% to 13% (p *<* 0.001), with median ICU stay rising from 4 to 7 days (p *<* 0.001). In-hospital mortality significantly decreased during 2022–2024 from 13.12% to 11.44% (p = 0.013). Mortality per 100,000 inhabitants was consistently higher in men.

**Conclusion:** This study highlights the high healthcare burden posed by acute embolism in hospitals, demonstrating an increase in patient complexity alongside a decline in overall mortality.

## Introduction

Acute pulmonary embolism (PE) is one of the main causes of cardiovascular morbidity, mortality, and hospitalization in developed countries [1]. Despite advances in diagnostic strategies and antithrombotic therapeutic regimens, the healthcare burden of this condition remains high, largely driven by an aging population and the prevalence of chronic comorbidities [2].

In the specific context of Spain, the epidemiological landscape has undergone significant changes over the last decade [3]. Analyzing historical trends is essential not only to understand the natural incidence of the disease, but also to evaluate the impact of disruptive events on the healthcare system. Studying patient‘s clinical complexity—assessed through tools such as the Charlson Comorbidity Index (CCI)—allows for better risk stratification and a deeper understanding of hospital outcome trends.

This study aims to describe the epidemiological characteristics and mortality of patients admitted for acute pulmonary embolism in Spanish hospitals between 2022 and 2024. Additionally, these findings are contextualized through a trend analysis dating back to 2016, providing a comprehensive view of the evolution of PE incidence and in-hospital case fatality rates within the Spanish health system.

## Methods

### Study Design, Population, and Data Collection

A nationwide, retrospective observational study was conducted to describe the epidemiological trends, clinical complexity, and in-hospital mortality of PE admissions in Spain from 2022 to 2024. Data were obtained from the Minimum Basic Data Set for Hospitalization (MBDS-H, or CMBD-H in Spanish) provided by the Spanish Ministry of Health.

### Selection of variables

Cases were identified by searching for the code I26 in any diagnostic position within the database. Several demographic and clinical variables were collected. Severity was calculated using the Charlson Comorbidity Index (CCI) to stratify baseline clinical complexity.

### Statistical Analysis

We summarized the main characteristics of the population using descriptive statistics, including median and interquartile range (IQR), as well as absolute values and percentages. To provide a standardized measure of disease burden, we calculated absolute values and rates per 100,000 population for hospitalization (HR) and mortality. Poisson regression models were used to assess the significance of trends over time for acute pulmonary embolism related admissions and mortality, with year treated as a continuous time variable. A p-value *<*0.05 was considered statistically significant. All analyses were conducted using R version 4.2.1.

### Ethical Considerations

The study was conducted in compliance with the Declaration of Helsinki. Data were obtained through a formal request to the Spanish Ministry of Health. Patient records were provided anonymized and de-identified, ensuring confidentiality and privacy. Because no names or personal information were recorded, no additional patient consent was required.

## Results

During the primary analysis period (2022–2024), a total of 125,305 admissions for acute PE were recorded in Spain, representing an HR of 0.89 per 100,000 population. The median age of patients remained constant at 74 years throughout the three-year period (p = 0.24). Regarding sex distribution, a slight male predominance was observed, with women representing 48–49A statistically significant increase in management complexity was identified: ICU admissions steadily rose from 11% in 2022 to 13% in 2024 (p *<* 0.001). The median ICU stay increased from 4 days (IQR: 2–9) in 2022 to 7 days (IQR: 2–22) in 2024 (p *<* 0.001). Regarding pre-existing conditions, the associated comorbidity burden was high, notably with a prevalence of COPD (Chronic Obstructive Pulmonary Disease) at 61–62%, hypertension at 35–37%, and malignancy at 21–24%. The Charlson Comorbidity Index (CCI) showed that over 36% of patients had a score *>* 4. Figure 1 shows trends in total admissions for acute pulmonary embolism in Spain from 2016 to 2022. Historical analysis reveals an upward trend in total admissions, rising from 27,577 in 2016 to a peak of 48,220 in 2021. Following this peak, annual incidence stabilized above 40,000 admissions per year during 2023–2024.

**Figure 1.**
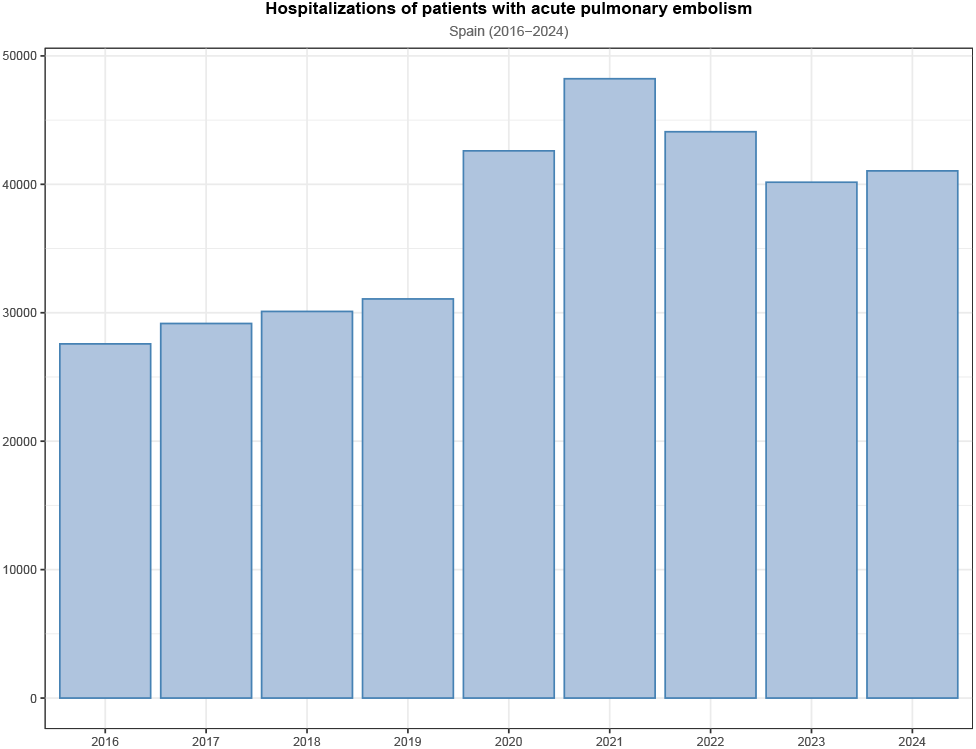
Hospitalization trend due to acute pulmonary embolism

Figure 2 shows mortality of acute pulmonary embolism. Regarding in-hospital mortality indicators, a significant reduction in the case fatality rate was observed during the primary study period, decreasing from 13.12% in 2022 to 11.44% in 2024 (p = 0.013). The highest in-hospital mortality rate was recorded in 2021 (13.84%) beginning in 2020.

**Figure 2.**
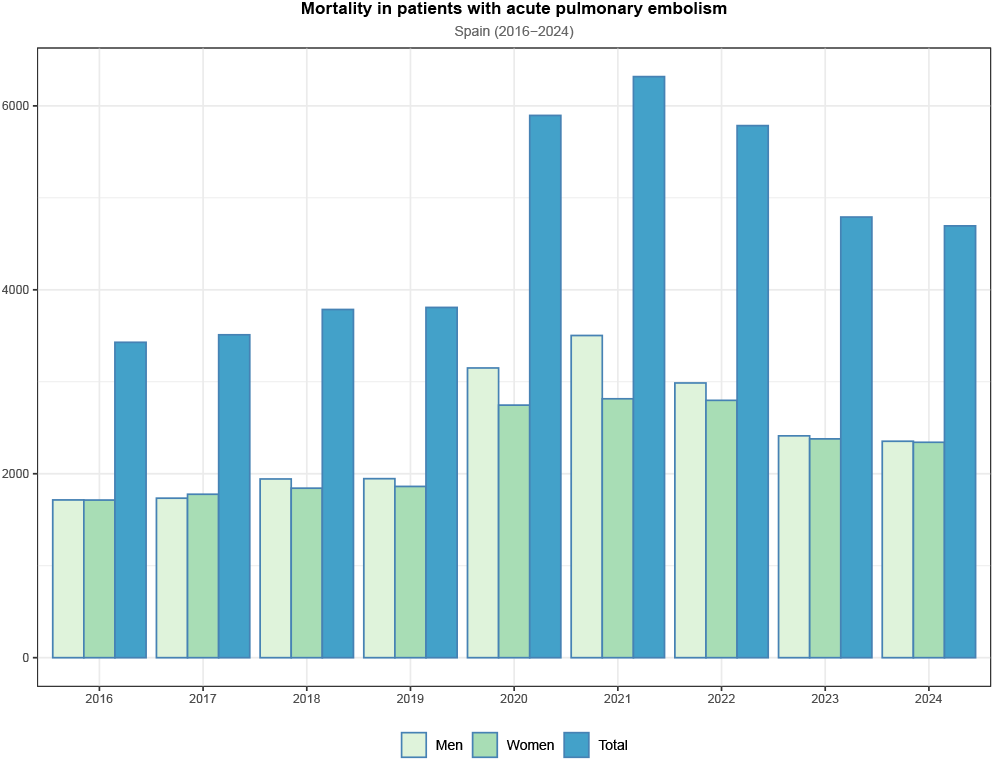
Mortality due to acute pulmonary embolism

Figure 3 shows mortality rate per 100,000 inhabitants for acute pulmonary embolism. Both sex groups saw a significant decline of mortality after the peak in 2021. When analyzing the rate per 100,000 inhabitants, men consistently exhibited higher mortality compared to women across the entire historical series. In 2021, the rate among men reached its highest point at approximately 15 deaths per 100,000 inhabitants, compared to 11.6 recorded among women.

**Figure 3.**
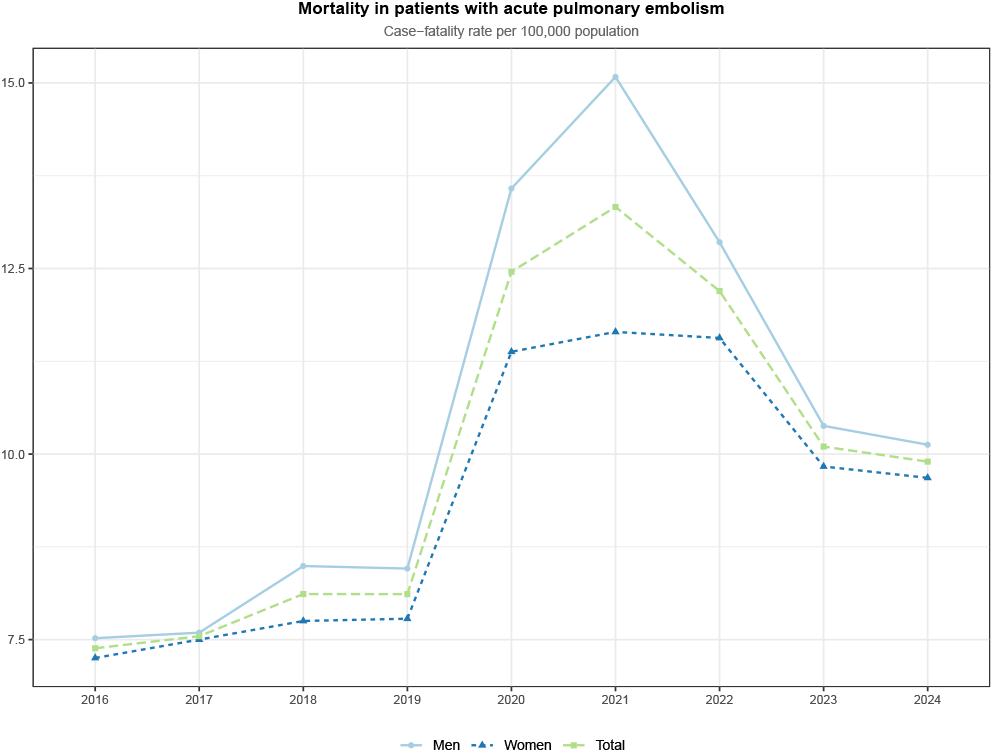
In-hospital mortality rate per 100,000 population due to pulmonary embolism

## Discussion

The descriptive analysis reveals a complex evolution marked by the impact of COVID-19 pandemic which was associated with substantially increased PE incidence and mortality [4]. Although outcomes improved by 2022, one of the most relevant findings is the stabilization of hospital admissions above 40,000 annually following the peak in 2021. Both incidence and mortality remained elevated compared to pre-pandemic era [5]. We observed a increase in the incidence of hospitalizations for pulmonary embolism, in line with the results of Hagiya et al. [6] in their analysis of data from the World Health Organization (WHO). Probably this increase is likely multifactorial, driven by an aging population and a higher prevalence of comorbidities that represent risk factors for thromboembolic diseases, such as cancer or obesity [7]. Despite this patient volume, a favorable trend in in-hospital mortality was observed, significantly decreasing from 13.12% to 11.44% over the last three-year period (p = 0.013). There are still not many studies [8] analyzing post-COVID-19 pandemic mortality, but this downward trend in pulmonary embolism mortality has already been observed in other studies. This decline could be attributed to improved early detection protocols—including optimized resource utilization through PE clinical probability scores, broader access to diagnostic imaging driven by recent technological advancements, and optimized therapeutic management. Indicators such as ICU length of stay and comorbidity burden, which usually correlate with disease severity, demonstrate an upward trend. The rise in ICU admissions (from 11% to 13%) and the prolonged median length of stay (from 4 to 7 days) suggest an increasing need for critical care and complex management. There are two factors that might account for the rise in both the number of ICU admissions and the length of stay. One is the fact that current patient profiles present a substantial burden of comorbidities, as mentioned earlier. The second is that the use of more advanced therapies, such as mechanical thrombectomy, has expanded in recent years [9], which often results in the patient requiring intensive post-procedural monitoring and specialized hemodynamic management. The results regarding mortality by sex are inconsistent, with some studies in line with our results, observing higher in-hospital mortality in men since others observed higher in-hospital mortality in women [10]. The causes of this phenomenon are still currently not fully understood by the scientific literature. Although admission volumes were similar between sexes, the mortality gap suggests the need to investigate potential sex-specific risk factors or differences in clinical presentation that might be influencing this outcomes.

## Conclusion

This study summarizes the epidemiological characteristics and analyzes the trends in the burden and mortality of pulmonary embolism in Spain in recent years, including the initial years following the COVID-19 pandemic. Incidence rates remain higher than pre-pandemic levels; however, despite treating more complex patients with longer ICU stays, a decrease in mortality is observed.

## Declarations

### Funding

No funding or sponsorship was received for this work.

### Authors’ contributions

Dr. Garcia-Carretero conceived and designed the study, wrote the first draft of the manuscript, and preprocessed and analyzed the data. Dr. Valle-Borrego, Dr. Peiro-Villalba and Dr. Martin-Rodrigo made substantial contributions to the interpretation of the results, critically reviewed the first draft of the manuscript, made valuable suggestions, and contributed to the visualization of the data. Drs. Quevedo-Soriano supervised the project and critically reviewed and edited the final draft of the manuscript. All authors read and approved the final manuscript.

### Availability of data and materials

Since the database was built upon open data, original datasets can be separately downloaded from their open sources. However, we have uploaded the compiled, final dataset that supports the findings of this study to a public repository, available at https://github.com/rafalinux/etev_2026.

### Ethics approval and consent to participate

No identifying information was included in the manuscript. Because the authors used historical data, informed consent was not necessary and no ethical approval was required. All procedures involving human participants were conducted in accordance with the ethical standards of the responsible institutional and/or national research committee and with the 1964 Helsinki Declaration and its later amendments or comparable ethical standards.

### Consent for publication

Not applicable.

### Competing interests

The authors have no conflicts of interest to declare.

### Declaration of generative AI and AI-assisted technologies in the manuscript preparation process

During the preparation of this work the authors did not use AI-assisted technologies. The authors take full responsibility for the content of the published article.

## Notes

### Competing Interest Statement

The authors have declared no competing interest.

### Author Declarations

Data were obtained through a formal request to the Spanish Ministry of Health. Patient records were provided anonymized and de-identified, ensuring confidentiality and privacy.

